# Percutaneous Coronary Intervention versus Optimal Medical Therapy on Quality of Life and Functional Capacity Assessment in Chronic Coronary Syndrome: A Meta-Analysis of Randomized Controlled Trials

**DOI:** 10.1101/2024.07.03.24309725

**Authors:** Sammudeen Ibrahim, Saint-Martin Allihien, Basilio Addo, Abdul-Fatawu Osman, Richard Amoateng, Haifz Sulemana, Jan Camille L. Ozaeta, Kwasi Asamoah Opare-Addo, Samuel Dadzie, Benedicta Arhinful, Shreyas Singireddy, Onoriode Kesiena, Sheriff N. Dodoo, Vedang Bhavsar

## Abstract

**Background:** Previous studies have primarily examined the impact of percutaneous coronary intervention (PCI) compared to optimal medical therapy (OMT) on hard outcomes such as all-cause mortality, cardiovascular death, nonfatal myocardial infarction, and the need for revascularization in patients with chronic coronary syndrome (CCS). However, these studies have not yielded significant findings thus far.

**Objective:** The goal of this meta-analysis was to assess the effect of PCI plus OMT on quality of life (QoL), functional capacity (FC), and angina-related health status compared to OMT alone in patients with CCS.

**Method:** Cochrane Central Registry of Controlled Trials, PubMed, Embase, and clininalTrials.gov were searched for studies published up to December 2023. The outcomes of interest were quality of life (QoL), freedom from angina (FFA), angina frequency (AF), and functional capacity (FC) measured with Seatle Angina Questionnaire (SAQ) or its equivalents such as EuroQol-5D (EQ-5D), 36-Item Short Form (SF-36) or RAND-36, and psychological well-being score if none is available. Additionally, the Duke Activity Status Index was also used to assess functional capacity when reported. Fixed-effect model was used for data analysis if I^2^ statistics <50%; otherwise, the random-effect model was used. Sensitivity analysis was performed by using the leave-one-out meta-analysis which alternatively removes a trial from the study to assess its impact on the result as well as the interconversion between fixed-effects and random-effects models. A meta-regression analysis was also performed to evaluate the impact of covariates on QoL. Cochrane Risk of Bias Assessment Tool was used to assess the risk of bias.

**Results:** Seventeen randomized controlled trials that enrolled 13,588 patients satisfied our inclusion and exclusion criteria with an average age of 62.3±10 years. PCI plus OMT improved QoL with standardized mean difference (SMD) of 0.27 ([95% CI, 0.14-0.40]; P <0.001) and 0.21 ([95% CI, 0.12-0.30]; P <0.001) when compared to OMT alone at 6 months and 1 year respectively. PCI plus OMT was also associated with significant improvement in FFA (1.17 [95% CI, 1.11-1.24]; P < 0.001), AF (0.25 [95% CI, 0.20-0.30]; P <0.001), and FC (0.21 [95% CI, 0.12-0.31]; P <0.001) at 6 months and persisted at 3 years (QoL [0.18 [95% CI, 0.06-0.30]; P < 0.001], FFA [1.27 [95% CI, 1.11-1.45]; P < 0.001], AF [0.09 [95% CI, 0.02-0.16]; P = 0.009], FC [0.13 [95% CI, 0.02-0.23]; P = 0.02] respectively) . However, there appears to be a reduction in this effect with time. Meta-regression analysis using year of publication as covariate on QoL was performed. There was no statistically significant relationship between the year of publication and QoL at 1 year (P = 0.81).

**Conclusion:** In this meta-analysis, PCI combined with OMT was associated with better quality of life, greater freedom from angina, reduction in angina frequency, and improved functional capacity compared to OMT alone. These benefits persisted when followed longitudinally for 3 years. However, longer-term outcome data of these trials is needed to determine whether these improvements in quality of life are sustained or attenuated over time.

## INTRODUCTION

Coronary artery disease (CAD) is the leading cause of death globally, accounting for 32% of global deaths in 2019^1^. The Center for Disease Control (CDC) reported that in the United States (US), 1 person dies from cardiovascular disease every 33 seconds^2^. CAD is therefore undeniably a matter of significant public health concern due to its widespread prevalence, substantial impact on morbidity and mortality, and considerable economic burden on healthcare systems globally^3^. Chronic coronary syndrome (CCS) represents a critical component of the CAD spectrum, characterized by a variable clinical course that may include chronic progression associated with periods of stability interspersed with unpredictable episodes of instability^3^. Although the benefit of percutaneous coronary intervention (PCI) in patients with acute coronary syndrome (ACS) on hard outcomes including all-cause death, cardiovascular death, and myocardial infarction has been well established, large randomized controlled trials (RCTs) found no benefit when compared to optimal medical therapy (OMT) in patients with CCS^4,5^. Thus the primary objective in managing patients with CCS has revolved around halting disease progression and enhancing patients’ overall health status which encompasses improving quality of life (QoL), functional capacity (FC), and addressing symptoms such as angina^6,7^. By pursuing these goals, healthcare providers may not only extend patients’ lifespan but also ensure that they can lead fulfilling and productive lives despite their condition.

The American Heart Association (AHA) recently recommended evaluating patient-reported QoL in patients with CCS^8^. Additionally, a recent study by Spertus et al., reported that invasive treatment strategies were associated with a significant improvement in quality of life compared to OMT alone^9^. Furthermore, a patient-level pooled analysis from the FAME 1 and 2 trials showed that PCI plus OMT was associated with improved quality of life in patients with higher angina class^10^ . Conversely, Al-Lamee and her colleagues reported no difference in quality of life when PCI plus OMT was compared to OMT plus sham procedure^11^. While the role of PCI in improving prognosis compared to OMT for patients with CCS remains a topic of debate, guidelines support its use to alleviate chronic angina and its overall impact on QoL that are not adequately managed with medical therapy alone^12,13^. As treatment options for CCS evolve, the primary therapeutic choices remain OMT alone or PCI combined with OMT^14^.

This systematic review and meta-analysis aimed to compare PCI plus OMT to OMT alone in patients with CCS. We evaluated both treatment strategies based on the QoL, angina frequency (AF), freedom from angina (FFA), and FC.

## 2 METHODS

This meta-analysis complied with the guidelines of Preferred Reporting Items for Systematic Reviews and Meta-analyses (PRISMA)^15^ and the Cochrane protocol^16^. Additionally, the study was registered under the International Prospective Register of Systematic Reviews (PROSPERO)^17^ with registration number *(CRD42024558472).* This study was exempted from institutional review board approval and informed consent of participants as the included studies were all publicly available, and their clinical data were de-identified in the individual trials ensuring compliance with ethical standards while conducting research involving human subjects.

### 2.1 Data Sources and Query Strategy

A comprehensive search was conducted across multiple databases, including PubMed, Cochrane Central Registry of Controlled Trials, Embase, and clinicalTrials.gov, to identify all relevant studies published up to December 2023. These trials assessed QoL, FFA, AF, and FC. The study evaluated validated measures such as the Short Form (SF)-36, RAND-36, Seattle Angina Questionnaire (SAQ), and Duke Activity Status Index (DASI)^18–21^. The quality of life was assessed using the SAQ, EuroQol-5D (EQ-5D), 36-Item Short Form (SF-36) or RAND-36, and psychological well-being score based on what the individual trial reported. The Duke Activity Status Index was used to assess functional capacity when reported. The SAQ evaluates four dimensions of angina-related quality of life: anginal symptom frequency, physical function/limitations, angina-specific quality of life, and treatment satisfaction. Additionally, the EQ-5D 0 to 100 visual analog scale was used to assess self-rated global health. The Duke Activity Status Index (DASI) to assess cardiac functional status, and the RAND General Health survey was used to obtain an overall health ordinal rating. These measures collectively provide a comprehensive understanding of patients’ well-being and functional status in the context of CCS. The comprehensive search protocol is available in Supplemental Material Table 1 for detailed reference.

**Table 1:**
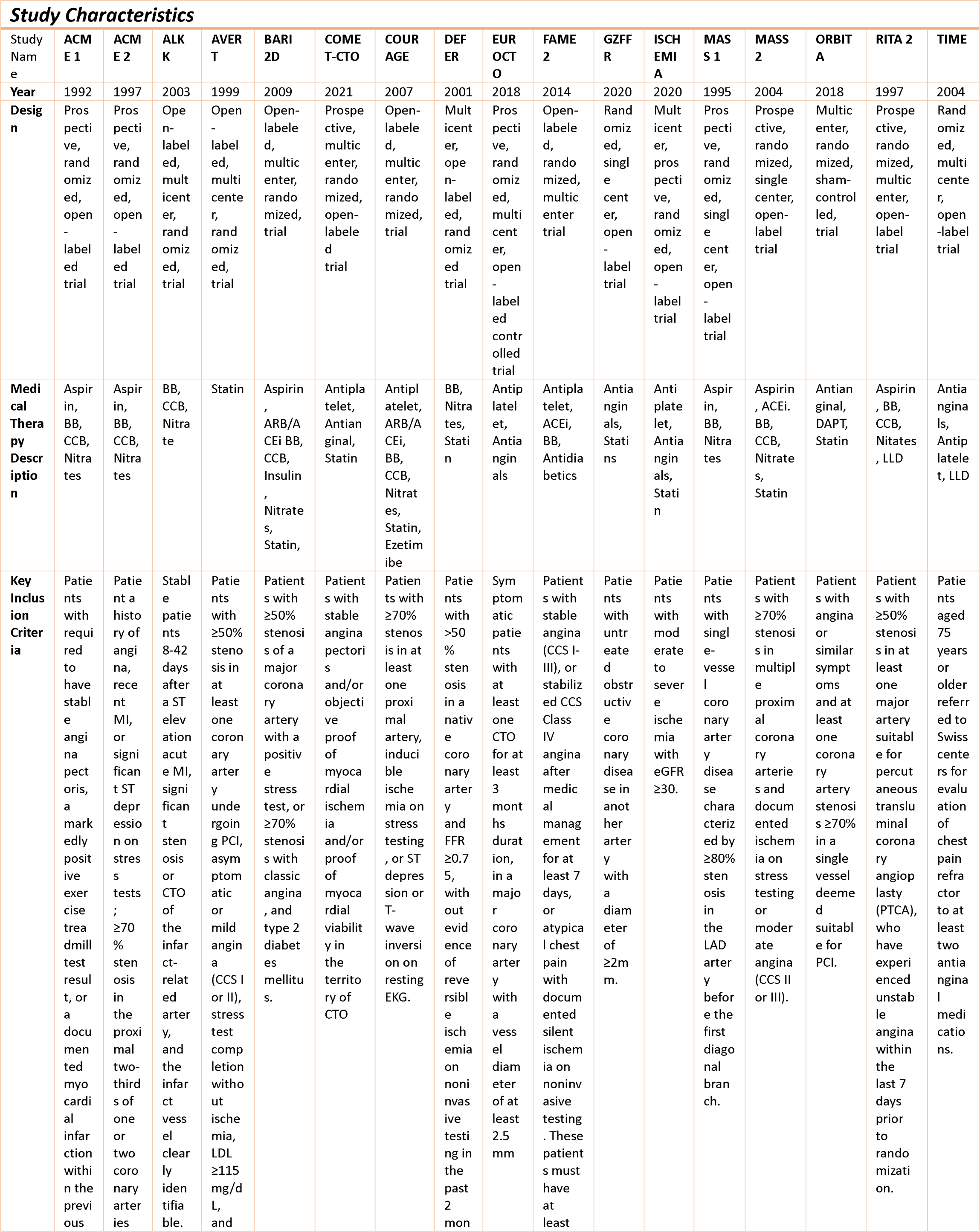
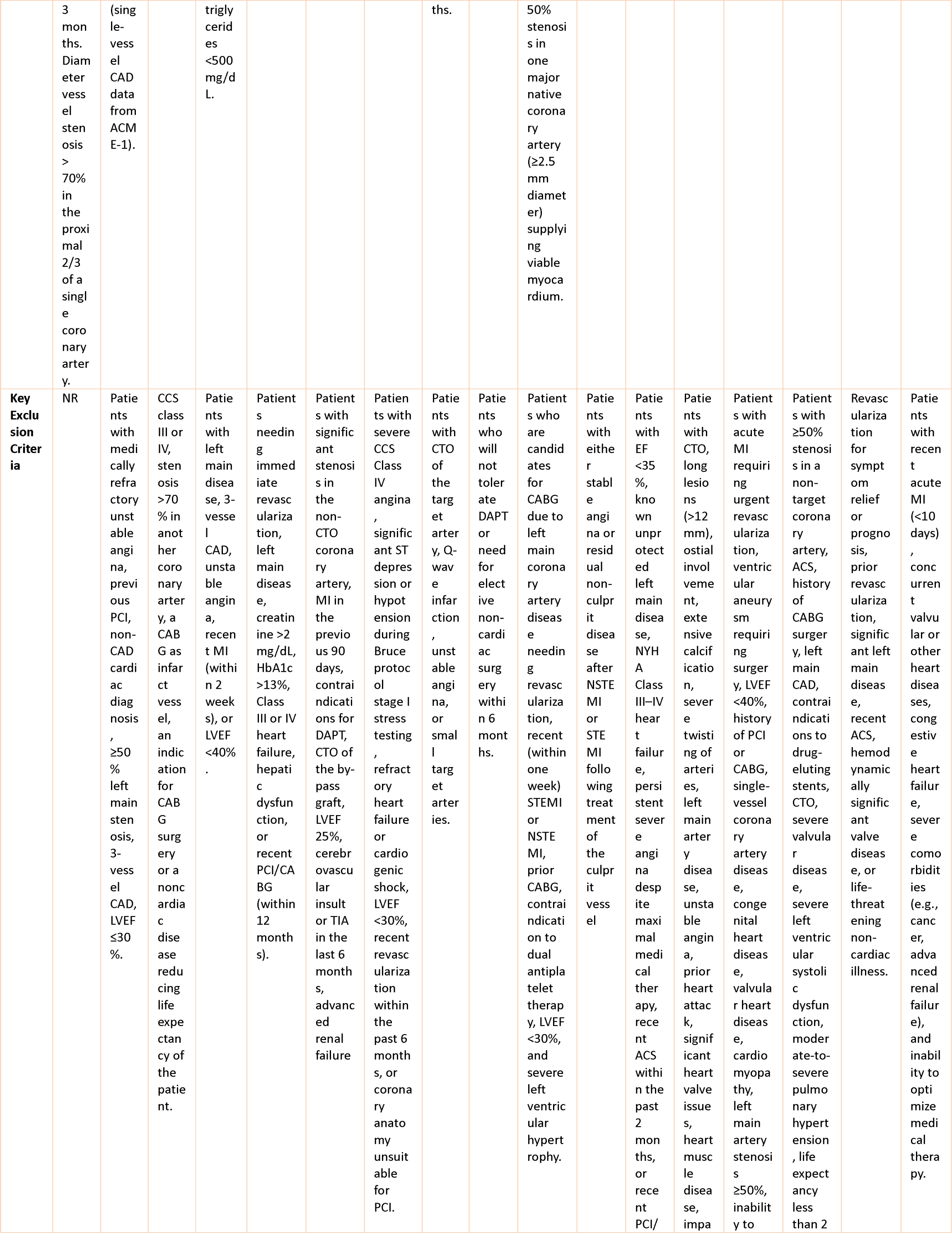
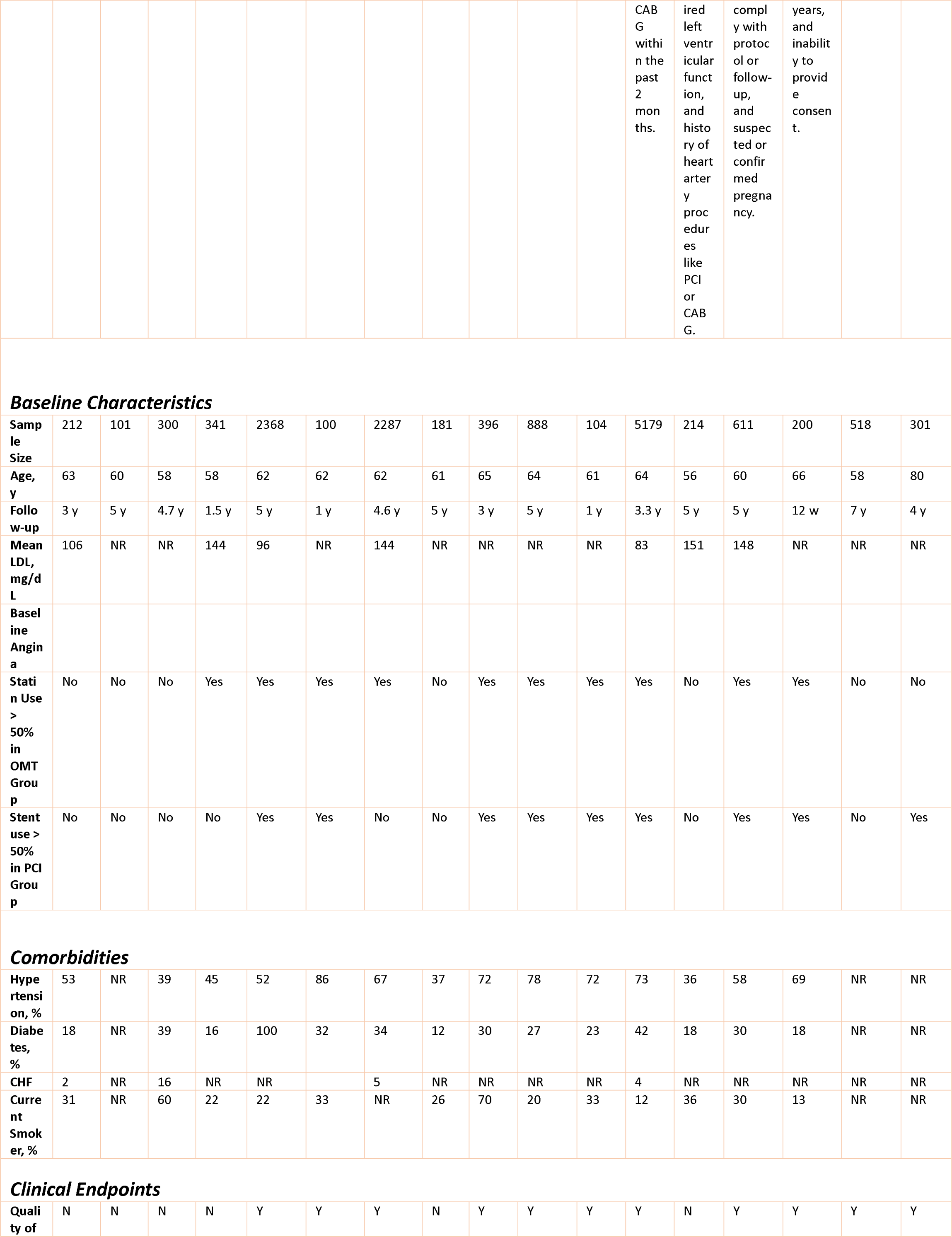
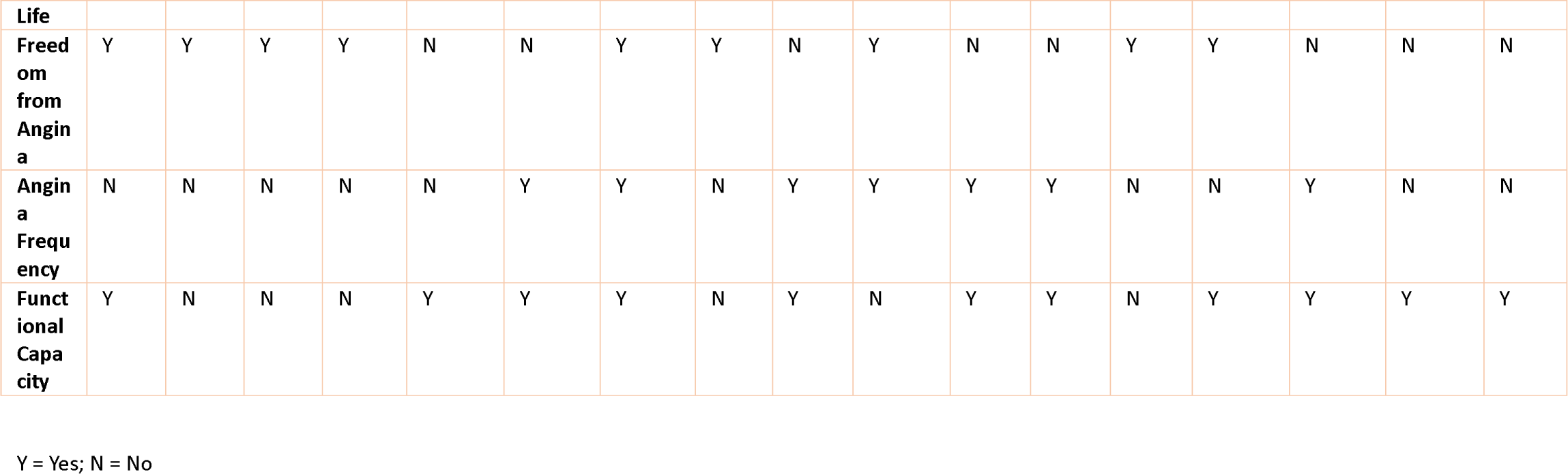
Select Study and Baseline Characteristics of Included Trials.

### 2.2 Study Selection

Two authors (SI and SA) independently screened all studies identified through the literature search, adhering strictly to predefined eligibility criteria derived from both inclusion and exclusion criteria. Any discrepancies between the two authors were resolved by a third author (OA). Ultimately, 17 RCTs that met the eligibility criteria were included in the final evaluation.

Eligible studies for inclusion comprised RCTs involving adult participants aged 18 years and above. These trials compared PCI plus OMT to OMT alone. We also included three-arm RCTs that compared PCI plus OMT versus OMT alone versus CABG in which the three comparative groups were clearly delineated and reported at least one outcome measures of interest. Exclusion criteria comprised studies lacking sufficient statistical data for comparison between the two treatment groups, those examining only one treatment modality, investigations conducted on non-human subjects, and ongoing trials lacking relevant published data. Additionally, case reports, reviews, editorials, commentaries, and conference abstracts were also excluded from the analysis.

### 2.3 Data Extraction

After identifying and removing all duplicated data, the relevant studies were exported to the Endnote Reference Manager (version x5: Clarivate Analytics). The prespecified variables from the selected studies were then incorporated into a dataset. This process was independently verified by another author (SND).

Study characteristics, including the first author, publication year, study design, sample size, patient population, key inclusion and exclusion criteria, and year of publication, were extracted. Additionally, baseline characteristics such as age, sex, race, LDL, medical therapy used, statin use, stent use, and comorbidities were extracted. The clinical endpoints of interest including QoL, FFA, AF, and FC were also extracted from the selected studies. These have been summarized in Table 1.

### 2.4 Included Studies and Quality Assessment

The PRISMA flow diagram depicting the selection process for studies included in the final analysis is presented in Supplemental Material Figure 1. A total of 17 RCTs^4,5,11,22–39^. Table 1 outlines the characteristics of these studies, while their quality assessment is depicted in Supplemental Materials Figure 2. The quality of the included studies was independently assessed by two authors (SI and SND) using the modified Cochrane Collaboration’s risk of bias tool for RCTs^40^. Additionally, publication bias was evaluated through visual inspection and funnel plot analysis, conducted independently by the same authors (SI and SND).

**Figure 1:**
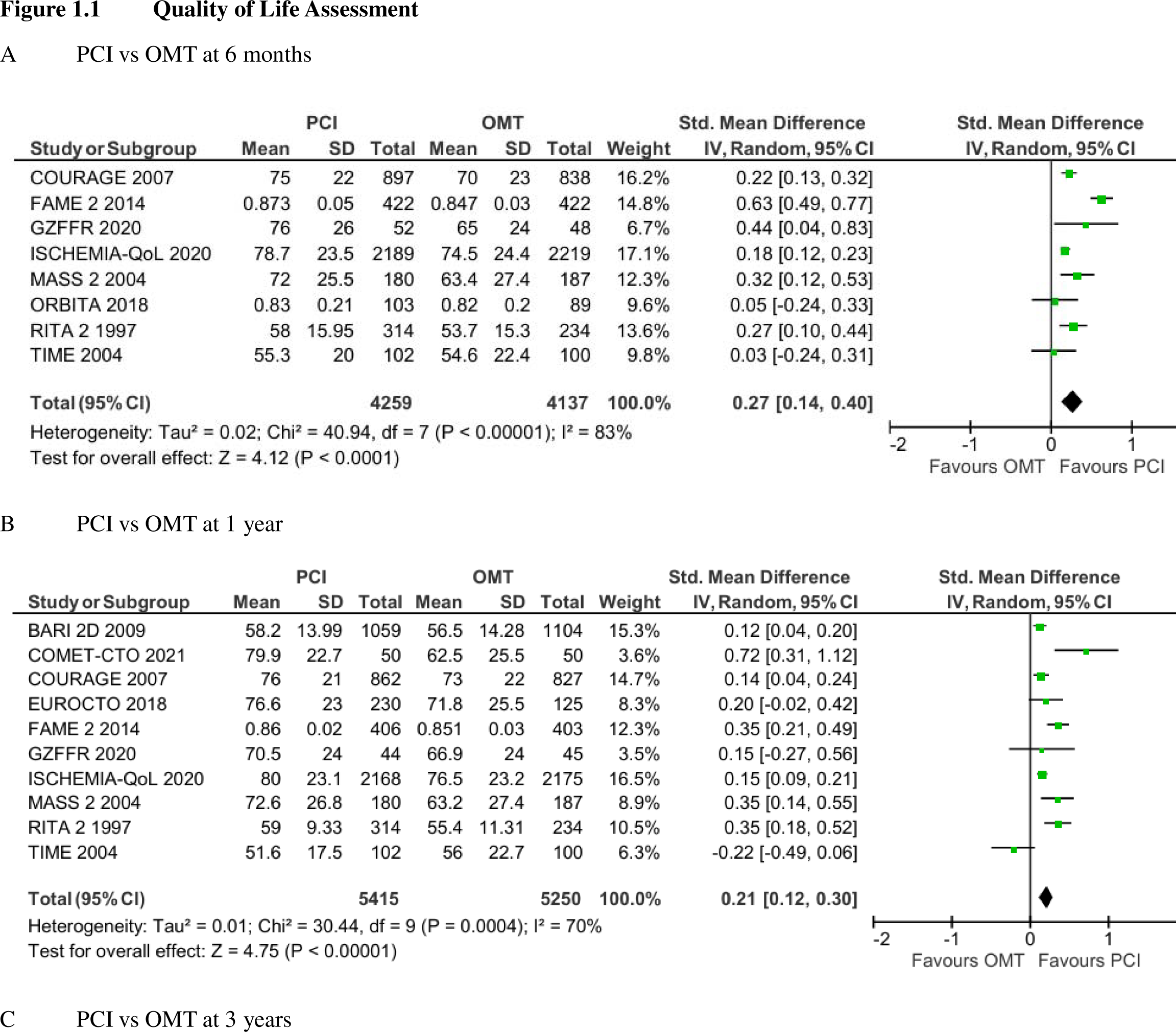
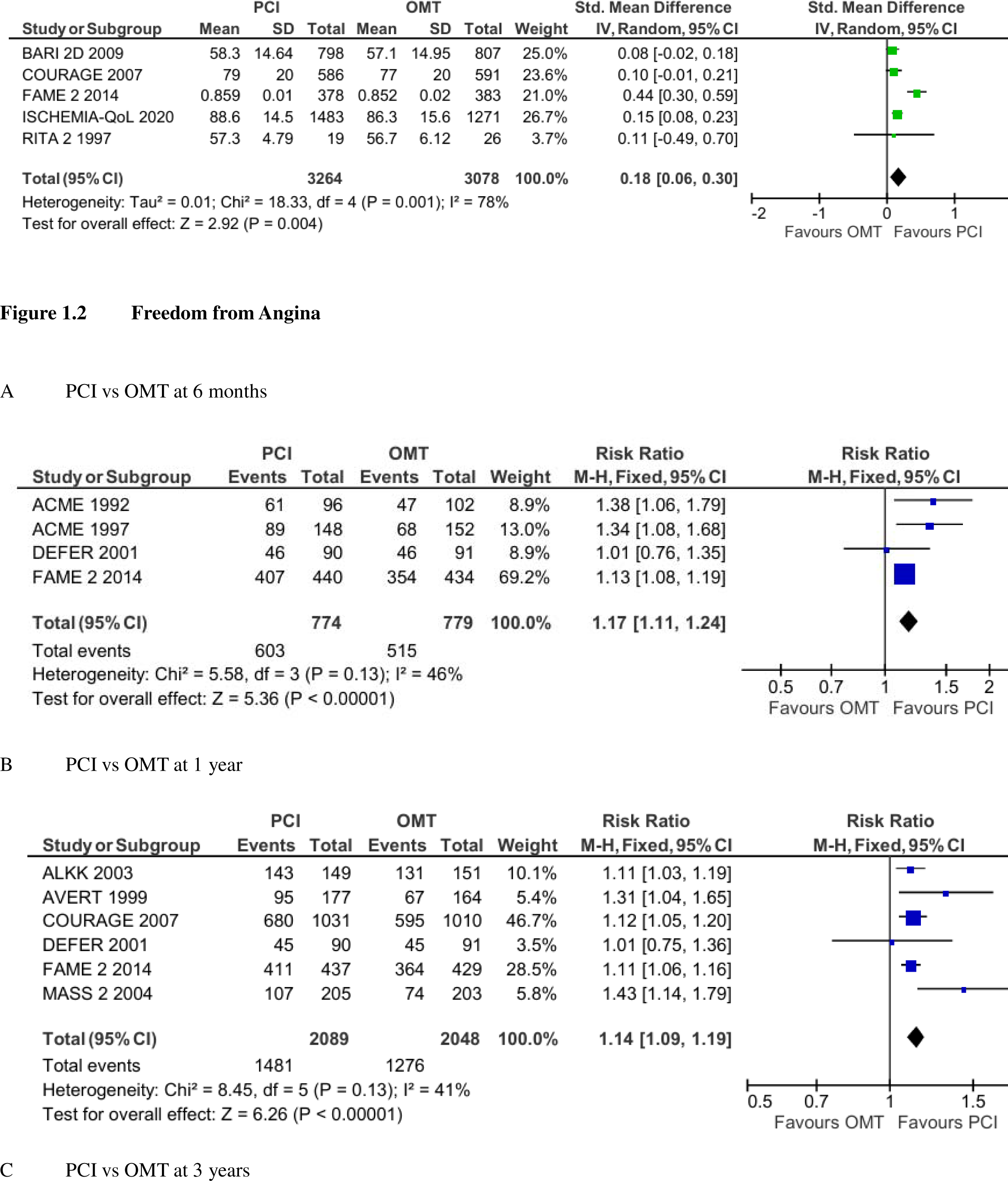
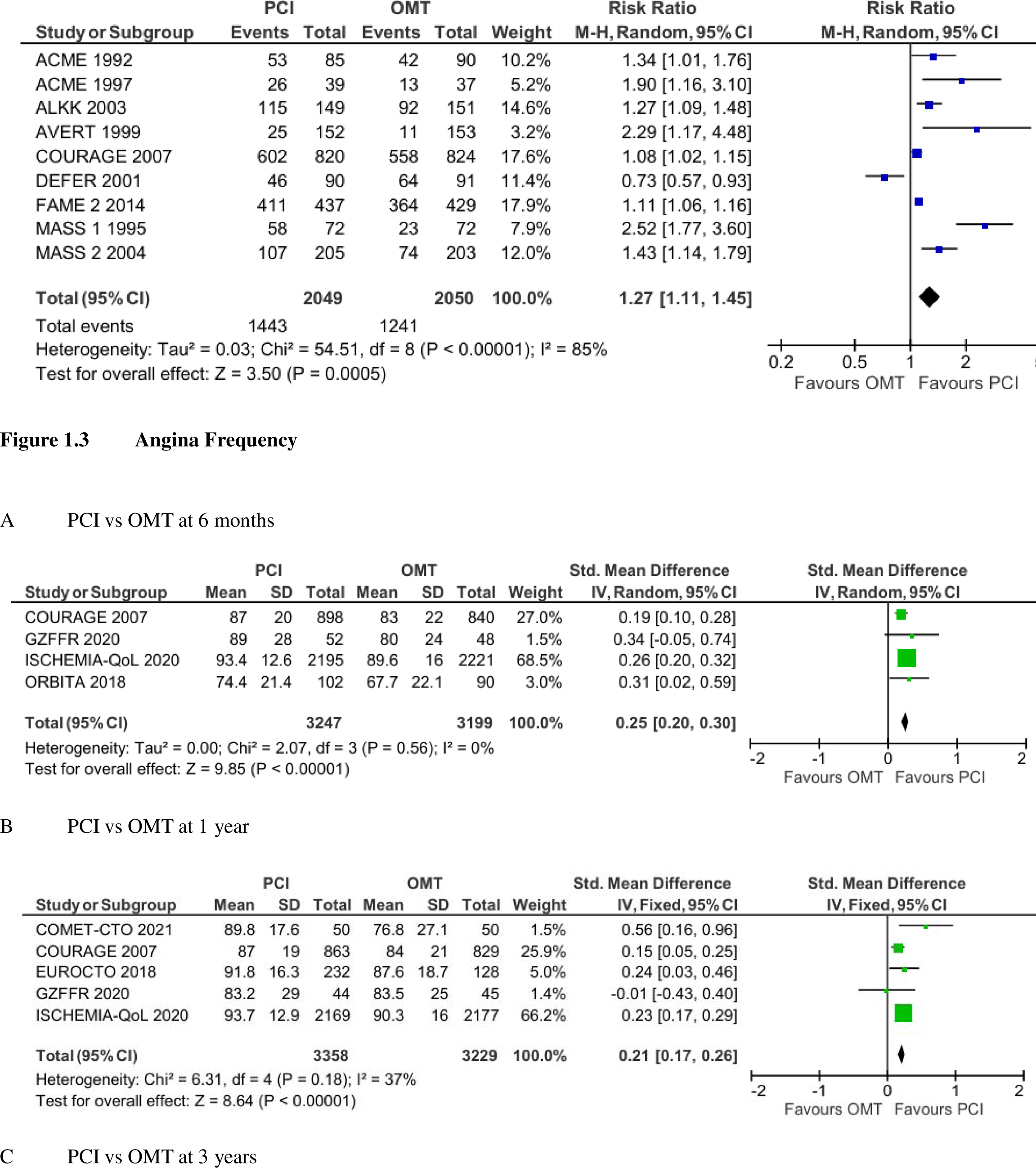
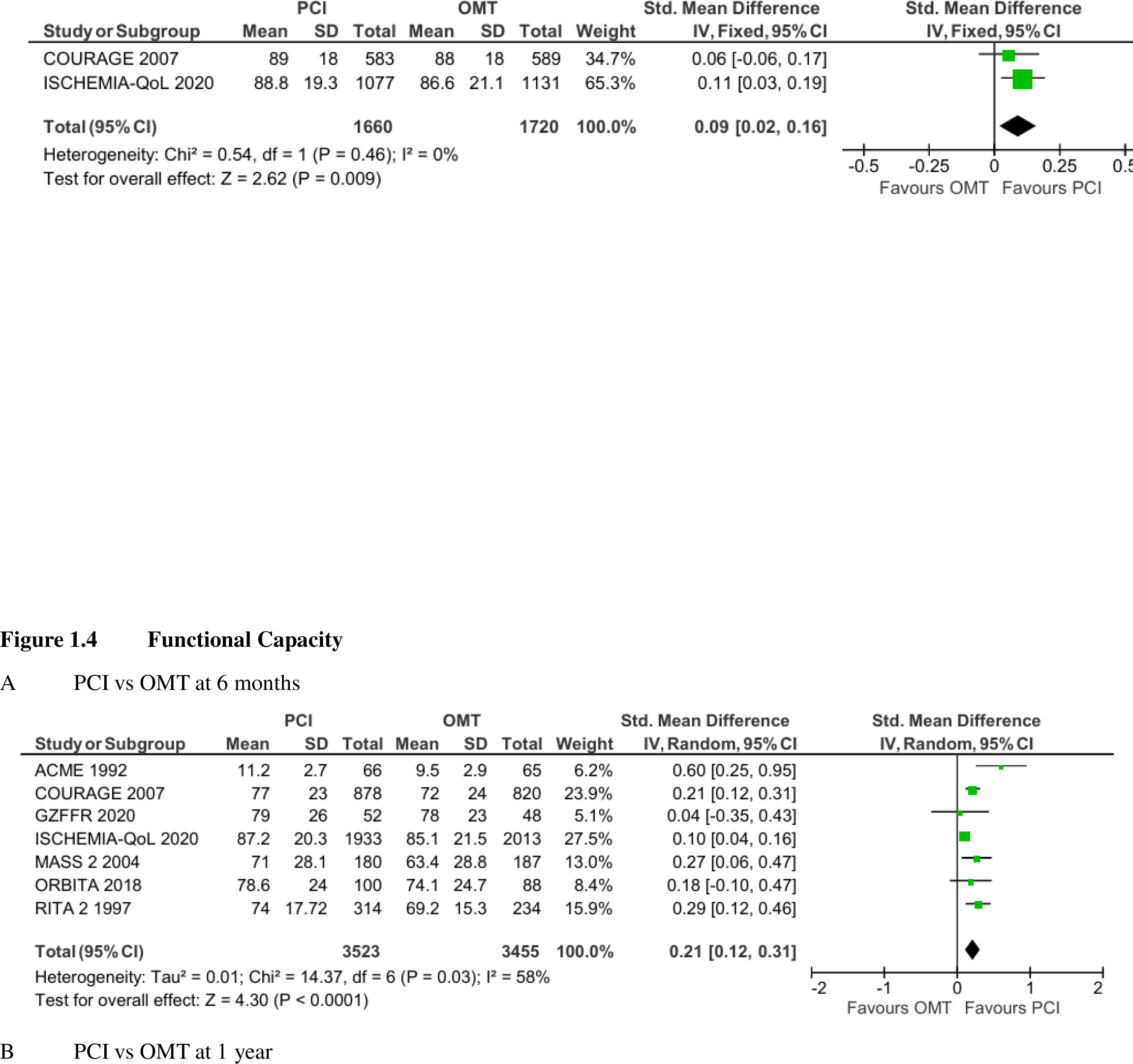
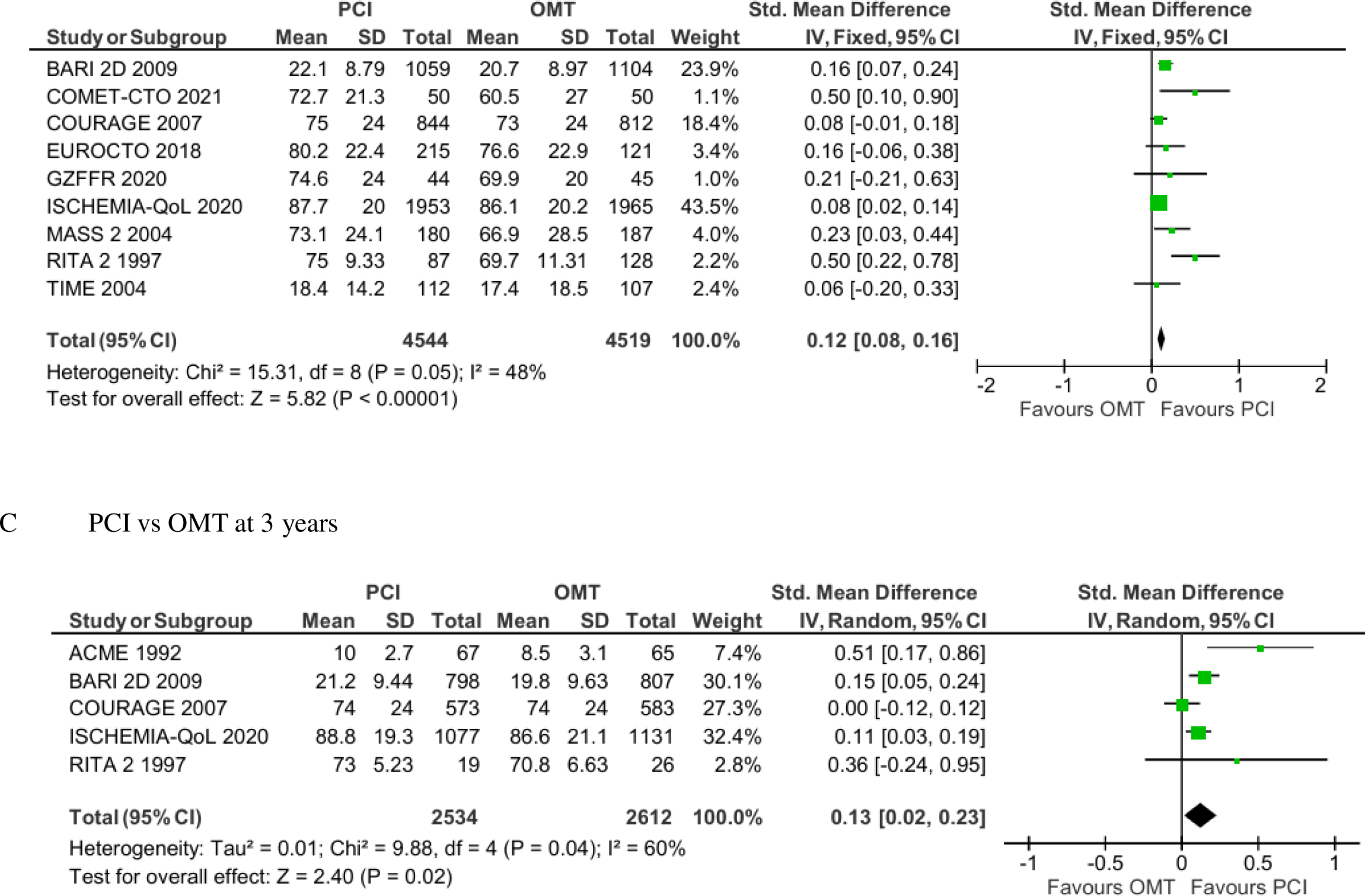
Showing the Forest Plot of Outcomes Comparing PCI plus OMT to OMT alone.

### 2.5 Statistical Analysis

All the statistical analyses were conducted using Review Manager (version 5.4.1; Copenhagen: The Nordic Cochrane Center, the Cochrane Collaboration, 2014) and Open Meta-Analyst. Mantel-Haenszel Summary statistics were presented using odds ratios (ORs) with corresponding 95% confidence intervals (CIs) for categorical variables. For continuous variables, standardized mean differences (SMDs) with 95% CIs were utilized. To assess heterogeneity between study protocols, Cochran’s Q and I^2^ statistics were employed. Heterogeneity was considered significant at a P value < 0.05 and an I^2^ value exceeding 50%. Depending on the degree of heterogeneity observed, either a fixed-effect model or a random-effect model was utilized to derive the pooled effect estimate.

To evaluate the robustness of the results of the meta-analysis, sensitivity analyses were performed using the leave-one-out analysis by systematically removing one trial at a time from the pool. This analysis allowed for an assessment of the impact of each individual trial on the overall result. Finally, sensitivity analysis was also conducted by interconversion between fixed-effects and random-effects models.

Analyses were stratified based on the use of fractional flow reserve (FFR) assessment prior to intervention and the inclusion of chronic total occlusions (CTOs) in culprit vessels in the studies. This stratification aimed to evaluate their impact on outcomes. We estimated the difference between the estimates of these subgroups using a test for interaction. Additionally, a meta-regression analysis was conducted to examine the relationship between the year of trial publication and QoL. This analysis was performed to determine whether the results differ in more recent trials in the setting of contemporary therapy modalities.

## 3. RESULTS

### 3.1 Characteristics of Study Participants

Seventeen RCTs including 13,588 patients were selected for the final analysis, with 6,899 in the PCI plus OMT arm and 6,689 in the OMT alone arm. All of the 17 RCTs were published between 1992 and 2022. The baseline characteristics of the participants of included RCTs and study inclusion/exclusion as well as clinical endpoints are reported in are summarized in Table 1. The included RCTs showed minimal publication bias, as depicted by the symmetrical funnel plot shown in Supplemental Material Figure 3. The included studies were all of sufficient quality as evident on the risk of bias assessment shown in Supplemental Material Figure 2.

### 3.2 Outcomes

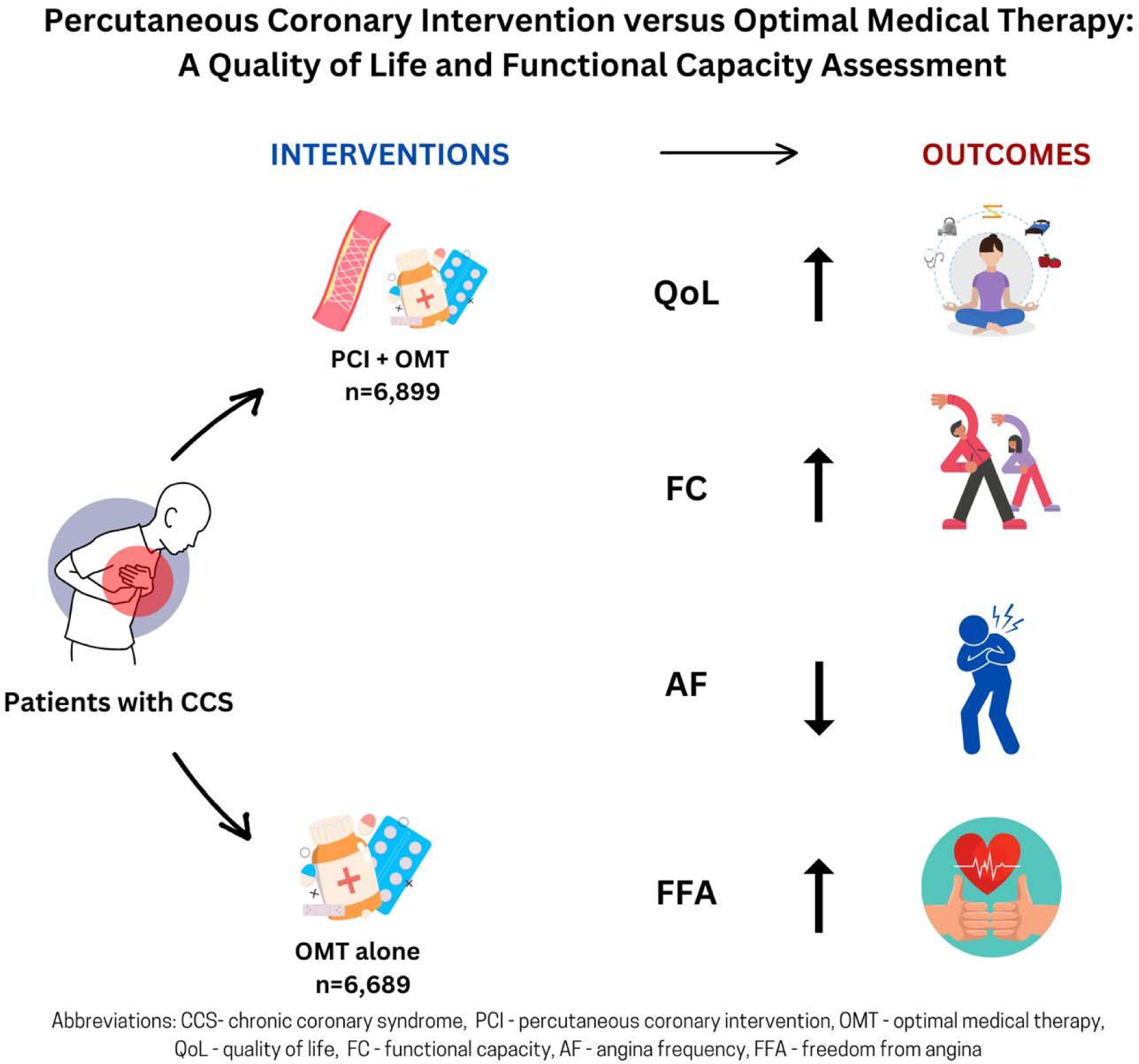

Central Illustration Summarizing Study and the Outcomes of Interest

#### 3.2.1 Quality of Life

Overall, 11 RCTs assessed QoL outcomes which comprised of 10,465 participants, with 5,346 in the PCI arm and 5,119 in the OMT arm. PCI was associated with improved quality of life when compared to OMT (SMD = 0.27 [95% CI, 0.14-0.40]; P <0.001), (SMD = 0.21 [95% CI, 0.12-0.30]; P <0.001), and (SMD = 0.18 [95% CI, 0.06-0.30]; P = 0.004) at 6 months, 1 year, and 3 or more years respectively. The results of the meta-analysis are shown in Figure 1.1.

#### 3.2.2 Freedom from Angina

A total of 9 RCTs reported on the effect of PCI compared to OMT on FFA including 4,787 participants with 2,408 and 2,379 in the PCI and OMT groups respectively. We study found that PCI was associated with significant increases in FFA when compared to OMT at 6 months (OR = 1.17 [95% CI, 1.11-1.24]; P < 0.001), 1 year (OR = 1.14 [95% CI, 1.09-1.19]; P <0.001), and 3 year or more (OR = 1.27 [95% CI, 1.11-1.45]; P = 0.004). The results of the meta-analysis are shown in Figure 1.2.

#### 3.2.3 Angina Frequency

Five RCTs evaluated the effects of PCI on AF when compared to angina frequency. This included a total of 6,546 patients with 3,297 in the PCI arm and 3,249 in the OMT arm. The effect estimate demonstrated a significant reduction in AF in favor of PCI at 6 months (SMD = 0.25 [95% CI, 0.20-0.30]; P <0.001), 1 year (SMD = 0.21 [95% CI, 0.17-0.26]; P < 0.001), and 3 or more years (SMD = 0.09 [95% CI, 0.02-0.16]; P = 0.009). The results of the meta-analysis are shown in Figure 1.3.

#### 3.2.4 Functional Capacity

Overall, 11 RCTs assessed FC including 9,796 participants with 4,959 in the PCI arm and 4,837 in the OMT arm. PCI was associated with a statistically significant improvement in FC when compared to OMT (SMD = 0.21 [95% CI, 0.12-0.31]; P < 0.001), (SMD = 0.12 [95% CI, 0.08-0.16]; P < 0.001), and (SMD = 0.13 [95% CI, 0.02-0.23]; P = 0.02) at 6 months, 1 year, and 3 or more years respectively. The results of the meta-analysis are shown in Figure 1.4.

### 3.3 Sensitivity Analysis

A sensitivity analysis was conducted using the leave-one-out method, where one trial was systematically removed from the pool at a time. This analysis allowed for an assessment of the impact of each individual trial on the overall results, identifying any single study that may disproportionately influence the pooled effect estimate. Additionally, sensitivity was further assessed by comparing the results obtained using both fixed-effects and random-effects models, allowing us to evaluate the robustness of the findings under different statistical assumptions. See Supplementary Material Figure 4.

### 3.4 Meta-regression Analysis

Meta-regression analyses were performed to investigate the relationship between the year of trial publication and the QoL outcomes over time. The results indicated no statistically significant association between the year of publication and QoL improvements, with a P value of 0.81. See Supplementary Material Figure 5.

## DISCUSSION

In this present meta-analysis of RCTs comparing PCI to OMT, we report several significant findings:

1. PCI was associated with improved QoL in patients with chronic coronary syndrome and persisted over 3 years.
2. FC, FFA, and AF were significantly improved by PCI when compared to OMT.
3. The effect of PCI on QoL, FC, FFA, and AF appears attenuated over time.

Numerous previous studies have indicated that in patients with CCS, PCI is not associated with a reduction in major adverse clinical events including death, recurrent myocardial infarction, and need for new revascularization procedures when compared to OMT^4,30,41,42^. Assessing soft outcomes including quality of life, functional capacity, angina-related health status has, therefore, gained significance in managing patients with CCS, a chronic condition known to impact quality of life and functional capacity^43,44^. Patient-reported outcome measures (PROMs) have been shown to play a crucial role in assessing the impact of healthcare services on the life of patients with CCS^45^. PROMs provide insights into health or well-being from the patient’s perspective, without clinician interpretation. According to the World Health Organization (WHO), health encompasses complete physical, mental, and social well-being, extending beyond the absence of disease or infirmity. Therefore, beyond extending life expectancy, patients prioritize the quality of additional life-years gained^46^. Consequently, understanding consistent symptomatic improvement following PCI is vital for physicians when discussing potential treatment benefits with patients.

The results of this current meta-analysis align with prior trials by showing improved quality of life of patients with CCS after PCI ^9,39,47,48^. The TIME Trial was one of the pioneering studies in stable CAD patients that demonstrated the benefit of invasive strategies over OMT in terms of quality of life^39^. Similarly, in the ISCHEMIA trial, patients with chronic coronary syndrome and moderate or severe ischemia on functional testing experienced clinically significant improvements in quality of life with an initial invasive strategy^9^. However, these benefits were observed primarily in patients with more frequent anginal episodes at the time of treatment evaluation^49^. The COURAGE trial also assessed quality of life changes between PCI plus OMT and OMT alone. Interestingly, while patients treated with PCI initially experienced improved quality of life, this effect diminished by the 12-month period^4^. Similarly, this current review also found that PCI was associated with better quality of life when compared to OMT and the effect size appeared to attenuate with time as well. Some studies have reported that the diminishing quality of life benefits observed over longer follow-up periods suggested that perhaps the initial short-term improvements from invasive approaches may be influenced, at least in part, by a placebo effect^4,5^. Indeed, in the first double-blinded, placebo-controlled trial using a sham procedure, ORBITA, Al-Lamee and her colleagues found no improvement in exercise time and quality of life scores beyond the sham procedure, even in patients with significant coronary stenosis^11^. Contrastingly, the findings from a more recent double-blinded, placebo-controlled trial, ORBITA-2, revealed that in patients with CCS who were receiving minimal or no antianginal medication and had an objective evidence of ischemia, PCI led to a reduced angina symptom score compared to a sham procedure^50^. This suggests an improvement in angina-related quality of life with PCI. However, it is important to note that even though the daily data showed that the effect of PCI was immediate and sustained, the patients in this study were followed for only 12 weeks, thus limiting the assessment of long-term effects beyond this timeframe.

Our current meta-analysis revealed that PCI was associated with improvements in angina-related health status, including freedom from angina and angina frequency, compared to OMT. Although this benefit persisted over a three-year follow-up period, we observed a decrease in the effect size over time. Consistent with our findings, a prior meta-analysis also demonstrated a significant improvement in angina-related health status with PCI plus OMT versus OMT alone. Notably, the inclusion of recent myocardial infarction (MI) survivors in some trials may have influenced these results^51^. Additionally, Boden et al also reported that the need for revascularization during follow-up was more common in patients receiving OMT, particularly in those with severe angina^4^. This trend aligns with previous studies indicating a decline in the proportion of angina-free patients with revascularization compared to initial medical therapy over time^52^. Although our analysis suggested a higher rate of revascularization in the OMT group, this difference did not reach statistical significance (See Supplemental Material 7). The attenuation in the magnitude of improvement in quality of life and angina-related health status over time may be partly attributed to high crossover rates from the OMT to PCI group. However, further research is needed to explore these findings and their implications for clinical practice.

Additionally, this meta-analysis demonstrated that PCI plus OMT led to improvements in functional capacity compared to OMT, as measured by various parameters including the DASI score, exercise tolerance testing, and the physical limitation component of the SAQ. Although this effect persisted over the three-year period, it appeared to diminish with time. Previous studies have also reported similar benefits of PCI plus OMT over OMT alone in functional capacity, with some indicating a more pronounced benefit in patients with lesions having lower baseline fractional flow reserve (FFR) values^11,22,24,30,50^. However, a subgroup analysis in this review found no significant impact of FFR on functional capacity. Another study focusing on physical activity after PCI reported sustained improvements in functional capacity up to 12 months, particularly in patients with chronic total occlusion (CTO)^55^. Nevertheless, a subgroup analysis in this current study found that the presence of CTO as the target vessel in the trial population did not influence the impact of PCI.

Indeed, while this current meta-analysis demonstrated that PCI in addition to OMT is superior to OMT alone in improving soft outcomes such as quality of life, functional capacity, angina relief, and reducing need for subsequent procedures, several important considerations remain unanswered. Firstly, it is uncertain whether the extent of angina reduction and the decrease in subsequent procedures achieved with PCI justify the increased costs associated with stenting and the procedural risks. Additionally, the potential periprocedural and long-term safety risks of PCI compared to OMT warrant careful consideration. The results of the cost-effectiveness analysis of PCI plus OMT versus OMT alone have varied across different trials. The TIME and FAME 2 trials indicated that PCI plus OMT was more cost-effective than OMT, suggesting favorable economic outcomes associated with the intervention^53,54^. However, analyses from trials such as COURAGE and BARI 2D reported contrasting findings, indicating that PCI plus OMT may not always be the more cost-effective option compared to OMT alone^55,56^. A comprehensive cost-effectiveness analysis revealed that the disparities observed in the cost-effectiveness of PCI plus OMT versus OMT alone may stem from the duration of follow-up^57^. Short-term studies may overestimate cost due to the initial high expenses associated with PCI, without adequately considering the long-term reduction in repeat procedures^57^. Moreover, the effectiveness of PCI in mitigating subsequent angina is notably contingent upon the initial symptom severity of the patient population, with PCI demonstrating greater benefits for individuals presenting with more severe symptoms^57^. These insights are crucial for informed decision-making and optimal management of patients with CCS.

## STUDY LIMITATIONS

We recognize several limitations to our analysis. Although the evaluation of quality of life, angina frequency, freedom from angina outcomes, and functional capacity were performed with validated questionnaires, the responses were all subjective and may be influenced by reporting bias from the participants. Similar to other trial-level analyses, we were unable to consider factors such as adherence to assigned treatment, duration of medical therapy, type of stent, or the proportion of patients with stent usage in our analysis. These factors would be best evaluated through an individual patient level meta-analysis. In addition to our sensitivity analysis on the impact of individual studies on the results we would have liked to explore the impact of optimal medical therapy (OMT) based on current guidelines. However, due to the significant heterogeneity from evolving nature of medical therapies and variations in blood pressure and cholesterol targets during the time of the individual trials, conducting such an analysis was not feasible. Furthermore, we may have underestimated the benefits of PCI plus OMT compared to OMT because several trials had a high crossover rate. Additionally, there was significant heterogeneity in the validated tool used to assess the clinical outcomes of interest. Lastly, our meta-regression analysis, which considered covariates such as year of publication in relation to quality of life, may be prone to ecological fallacy since we did not have access to individual patient data.

## CONCLUSION

The findings from our current meta-analysis demonstrate that PCI yields several benefits for patients with CCS over a period of 3 years compared to an initial approach of OMT. Specifically, PCI was associated with improvements in quality of life, functional capacity, freedom from angina, and reduction in angina frequency. Although there was a trend towards a reduced need for revascularization with PCI, this difference did not reach statistical significance. These compelling results highlight the potential long-term advantages of PCI in managing CCS, offering valuable insights for clinical practice and patient care.

## Data Availability

All data produced in the present study are available upon reasonable request to the authors.

---

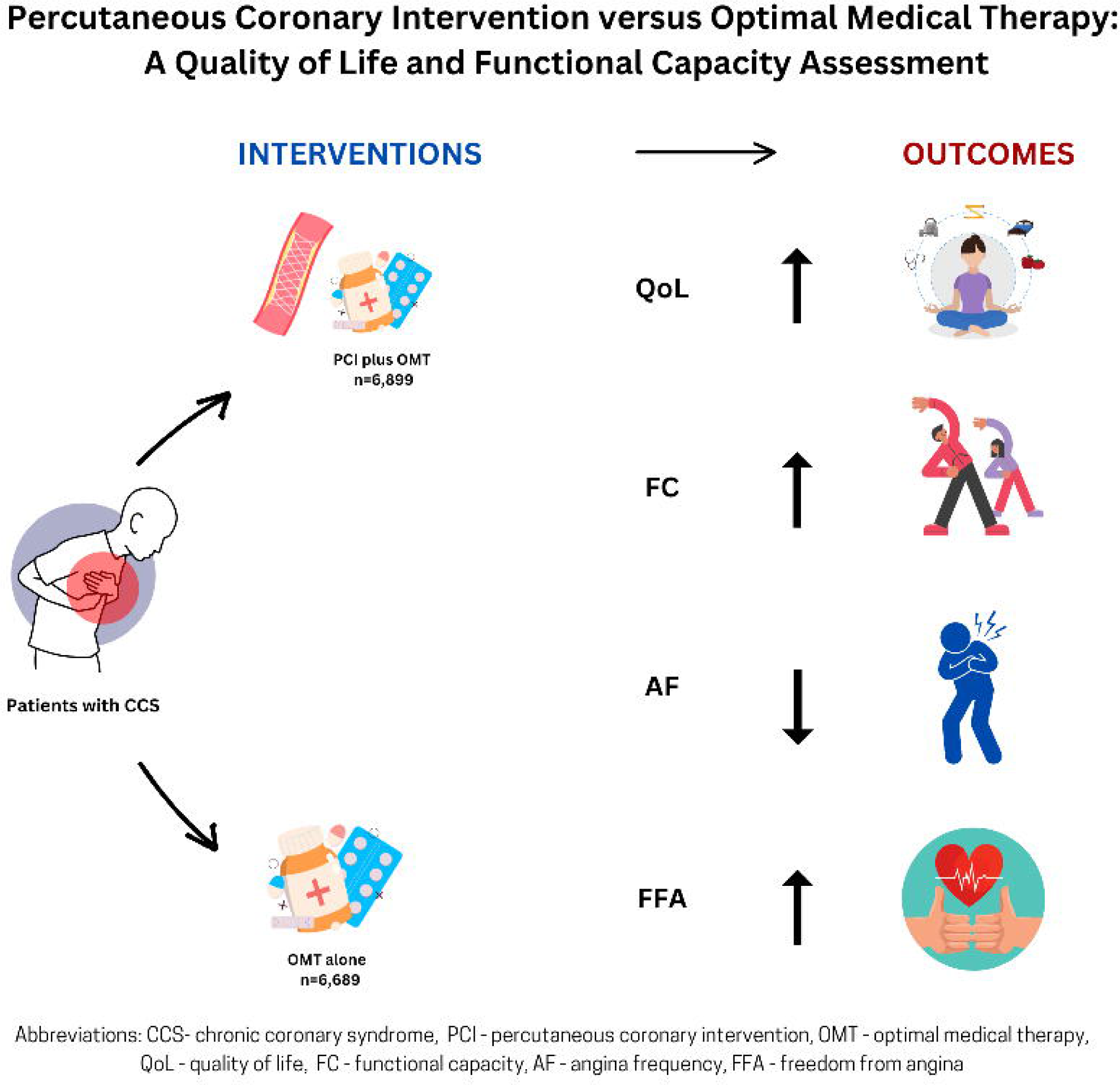

