## Supplemental Material for "Percutaneous Coronary Intervention versus Optimal Medical Therapy on Quality of Life and Functional Capacity Assessment in Chronic Coronary Syndrome: A Meta-Analysis of Randomized Controlled Trials"

| *Study Characteristics* | | | | | | | | | | | | | | | | | |
| --- | --- | --- | --- | --- | --- | --- | --- | --- | --- | --- | --- | --- | --- | --- | --- | --- | --- |
| Study Name | **ACME 1** | **ACME 2** | **ALKK** | **AVERT** | **BARI 2D** | **COMET-CTO** | **COURAGE** | **DEFER** | **EUROCTO** | **FAME 2** | **GZFFR** | **ISCHEMIA** | **MASS 1** | **MASS 2** | **ORBITA** | **RITA 2** | **TIME** |
| Year | 1992 | 1997 | 2003 | 1999 | 2009 | 2021 | 2007 | 2001 | 2018 | 2014 | 2020 | 2020 | 1995 | 2004 | 2018 | 1997 | 2004 |
| Design | Prospective,  randomized,  open-labeled  trial | Prospective,  randomized,  open-labeled  trial | Open-labeled, multicenter, randomized, trial | Open-labeled, multicenter, randomized, trial | Open-labeled, multicenter, randomized, trial | Prospective, multicenter,  randomized,  open-labeled  trial | Open-labeled, multicenter, randomized, trial | Multicenter,  open-labeled, randomized  trial | Prospective, randomized, multicenter, open-labeled controlled trial | Open-labeled, randomized, multicenter trial | Randomized, single center, open-label trial | Multicenter, prospective, randomized, open-label trial | Prospective, randomized, single center, open-label trial | Prospective, randomized, single center, open-label trial | Multicenter, randomized, sham-controlled, trial | Prospective, randomized, multicenter, open-label trial | Randomized, multicenter, open-label trial |
| Medical Therapy Description | Aspirin, BB, CCB, Nitrates | Aspirin, BB, CCB, Nitrates | BB, CCB, Nitrate | Statin | Aspirin, ARB/ACEi BB, CCB, Insulin, Nitrates, Statin, | Antiplatelet, Antianginal, Statin | Antiplatelet, ARB/ACEi, BB, CCB, Nitrates, Statin, Ezetimibe | BB, Nitrates, Statin | Antiplatelet, Antianginals | Antiplatelet, ACEi, BB, Antidiabetics | Antianginals, Statins | Antiplatelet, Antianginals, Statin | Aspirin, BB, Nitrates | Aspirin, ACEi. BB, CCB, Nitrates, Statin | Antianginal, DAPT, Statin | Aspirin, BB, CCB, Nitates, LLD | Antianginals, Antiplatelet, LLD |
| Key Inclusion Criteria | Patients with  required to have stable angina pectoris, a markedly  positive exercise treadmill test result, or a documented  myocardial infarction within the previous 3 months. Diameter  vessel stenosis > 70% in the proximal 2/3 of a single  coronary artery. | Patient a history of angina, recent MI, or significant ST depression on stress tests; ≥70% stenosis in the proximal two-thirds of one or two coronary arteries (single-vessel CAD data from ACME-1). | Stable patients 8-42 days after a ST elevation acute MI, significant stenosis or CTO of the  infarct-related artery, and the infarct vessel clearly identifiable. | Patients with ≥50% stenosis in at least one coronary artery undergoing PCI, asymptomatic or mild angina (CCS I or II), stress test completion without ischemia, LDL ≥115 mg/dL, and triglycerides <500 mg/dL. | Patients with ≥50% stenosis of a major coronary artery with a positive stress test, or ≥70% stenosis with classic angina, and type 2 diabetes mellitus. | Patients with stable angina pectoris and/or objective proof of myocardial ischemia and/or  proof of myocardial viability in the territory of CTO | Patients with ≥70% stenosis in at least one proximal artery, inducible ischemia on stress testing, or ST depression or T-wave inversion on resting EKG. | Patients with >50% stenosis in a native coronary artery and FFR ≥0.75, without evidence of reversible ischemia on noninvasive testing in the past 2 months. | Symptomatic patients with at least one CTO for at least 3 months duration, in a major coronary artery with a vessel diameter of at least 2.5 mm | Patients with stable angina (CCS I-III), or stabilized CCS Class IV angina after medical management for at least 7 days, or atypical chest pain with documented silent ischemia on noninvasive testing. These patients must have at least 50% stenosis in one major native coronary artery (≥2.5 mm diameter) supplying viable myocardium. | Patients with untreated obstructive coronary disease in another artery with a diameter of ≥2mm. | Patients with moderate to severe ischemia with eGFR ≥30. | Patients with single-vessel coronary artery disease characterized by ≥80% stenosis in the LAD artery before the first diagonal branch. | Patients with ≥70% stenosis in multiple proximal coronary arteries and documented ischemia on stress testing or moderate angina (CCS II or III). | Patients with angina or similar symptoms and at least one coronary artery stenosis ≥70% in a single vessel deemed suitable for PCI. | Patients with ≥50% stenosis in at least one major artery suitable for percutaneous transluminal coronary angioplasty (PTCA), who have experienced unstable angina within the last 7 days prior to randomization. | Patients aged 75 years or older referred to Swiss centers for evaluation of chest pain refractor to at least two antianginal medications. |
| Key Exclusion Criteria | NR | Patients with medically refractory unstable angina, previous PCI, non-CAD cardiac diagnosis, ≥50% left main stenosis, 3-vessel CAD, LVEF ≤30%. | CCS class III or IV, stenosis >70% in another coronary artery, a CABG as infarct vessel, an indication for CABG surgery or a noncardiac disease reducing life expectancy of the patient. | Patients with left main disease, 3-vessel CAD, unstable angina, recent MI (within 2 weeks), or LVEF <40%. | Patients needing immediate revascularization, left main disease, creatinine >2 mg/dL, HbA1c >13%, Class III or IV heart failure, hepatic dysfunction, or recent PCI/CABG (within 12 months). | Patients with significant stenosis in the non-CTO coronary artery, MI in the previous 90 days, contraindications for DAPT, CTO of the by-  pass graft, LVEF 25%, cerebrovascular insult or TIA in the last 6 months, advanced renal failure | Patients with severe CCS Class IV angina, significant ST depression or hypotension during Bruce protocol stage I stress testing, refractory heart failure or cardiogenic shock, LVEF <30%, recent revascularization within the past 6 months, or coronary anatomy unsuitable for PCI. | Patients with CTO of the target artery, Q-wave infarction, unstable angina, or small target arteries. | Patients who will not tolerate DAPT or need for elective non-cardiac surgery within 6 months. | Patients who are candidates for CABG due to left main coronary artery disease needing revascularization, recent (within one week) STEMI or NSTEMI, prior CABG, contraindication to dual antiplatelet therapy, LVEF <30%, and severe left ventricular hypertrophy. | Patients with either stable angina or residual non-culprit disease after NSTEMI or STEMI following treatment of the culprit vessel | Patients with EF <35%, known unprotected left main disease, NYHA Class III–IV heart failure, persistent severe angina despite maximal medical therapy, recent ACS within the past 2 months, or recent PCI/CABG within the past 2 months. | Patients with CTO, long lesions (>12 mm), ostial involvement, extensive calcification, severe twisting of arteries, left main artery disease, unstable angina, prior heart attack, significant heart valve issues, heart muscle disease, impaired left ventricular function, and history of heart artery procedures like PCI or CABG. | Patients with acute MI requiring urgent revascularization, ventricular aneurysm requiring surgery, LVEF <40%, history of PCI or CABG, single-vessel coronary artery disease, congenital heart disease, valvular heart disease, cardiomyopathy, left main artery stenosis ≥50%, inability to comply with protocol or follow-up, and suspected or confirmed pregnancy. | Patients with ≥50% stenosis in a non-target coronary artery, ACS, history of CABG surgery, left main CAD, contraindications to drug-eluting stents, CTO, severe valvular disease, severe left ventricular systolic dysfunction, moderate-to-severe pulmonary hypertension, life expectancy less than 2 years, and inability to provide consent. | Revascularization for symptom relief or prognosis, prior revascularization, significant left main disease, recent ACS, hemodynamically significant valve disease, or life-threatening non-cardiac illness. | Patients with recent acute MI (<10 days), concurrent valvular or other heart diseases, congestive heart failure, severe comorbidities (e.g., cancer, advanced renal failure), and inability to optimize medical therapy. |
| *Baseline Characteristics* | | | | | | | | | | | | | | | | | |
| Sample Size | 212 | 101 | 300 | 341 | 2368 | 100 | 2287 | 181 | 396 | 888 | 104 | 5179 | 214 | 611 | 200 | 518 | 301 |
| Age, y | 63 | 60 | 58 | 58 | 62 | 62 | 62 | 61 | 65 | 64 | 61 | 64 | 56 | 60 | 66 | 58 | 80 |
| Follow-up | 3 y | 5 y | 4.7 y | 1.5 y | 5 y | 1 y | 4.6 y | 5 y | 3 y | 5 y | 1 y | 3.3 y | 5 y | 5 y | 12 w | 7 y | 4 y |
| Mean LDL, mg/dL | 106 | NR | NR | 144 | 96 | NR | 144 | NR | NR | NR | NR | 83 | 151 | 148 | NR | NR | NR |
| Baseline Angina |  |  |  |  |  |  |  |  |  |  |  |  |  |  |  |  |  |
| Statin Use > 50% in OMT Group | No | No | No | Yes | Yes | Yes | Yes | No | Yes | Yes | Yes | Yes | No | Yes | Yes | No | No |
| Stent use > 50% in PCI Group | No | No | No | No | Yes | Yes | No | No | Yes | Yes | Yes | Yes | No | Yes | Yes | No | Yes |
| *Comorbidities* | | | | | | | | | | | | | | | | | |
| Hypertension, % | 53 | NR | 39 | 45 | 52 | 86 | 67 | 37 | 72 | 78 | 72 | 73 | 36 | 58 | 69 | NR | NR |
| Diabetes, % | 18 | NR | 39 | 16 | 100 | 32 | 34 | 12 | 30 | 27 | 23 | 42 | 18 | 30 | 18 | NR | NR |
| CHF | 2 | NR | 16 | NR | NR |  | 5 | NR | NR | NR | NR | 4 | NR | NR | NR | NR | NR |
| Current Smoker, % | 31 | NR | 60 | 22 | 22 | 33 | NR | 26 | 70 | 20 | 33 | 12 | 36 | 30 | 13 | NR | NR |
| *Clinical Endpoints* | | | | | | | | | | | | | | | | | |
| Quality of Life | N | N | N | N | Y | Y | Y | N | Y | Y | Y | Y | N | Y | Y | Y | Y |
| Freedom from Angina | Y | Y | Y | Y | N | N | Y | Y | N | Y | N | N | Y | Y | N | N | N |
| Angina Frequency | N | N | N | N | N | Y | Y | N | Y | Y | Y | Y | N | N | Y | N | N |
| Functional Capacity | Y | N | N | N | Y | Y | Y | N | Y | N | Y | Y | N | Y | Y | Y | Y |

Y = Yes; N = No; NR = Not reported; PCI = Percutaneous coronary intervention; OMT = Optimal medical therapy; CABG = Coronary artery bypass graft
